# A Multicenter Pilot Study of MUC1 Vaccine in Current and Former Smokers at High Risk for Lung Cancer

**DOI:** 10.1101/2025.06.25.25328823

**Authors:** Arjun Pennathur, David Midthun, Malgorzata Wojtowicz, Julie Ward, Judy Forster, Tami Krpata, Samantha Fatis, John McKolanis, Jia Xue, Pamela Beatty, Sharon Kaufman, Colleen Akerley, April Felt, Karrie Fursa, Anne Holland, Liz Ambulay, Nathan R. Foster, Carrie Strand, Andres Salazar, Lisa Bengtson, Eva Szabo, Paul Limburg, Olivera J. Finn

**Author notes:** Corresponding Authors: Olivera J. Finn, Ph.D., University of Pittsburgh School of Medicine, Department of Immunology, The Assembly Building, 5051 Centre Ave, Pittsburgh, PA 15213; and Arjun Pennathur MD, Department of Cardiothoracic Surgery, University of Pittsburgh School of Medicine, 200 Lothrop St Suite C-800, Pittsburgh, PA 15213.

## Abstract

**Background:** Smoking is the most common etiology for lung cancer and smoking cessation does not eliminate the risk. Mucin (MUC)1 glycoprotein is aberrantly expressed in lung carcinomas and premalignant lung lesions. We explored whether a MUC1 vaccine might be effective in halting neoplastic development and progression in individuals at high risk for lung cancer.

**Methods:** We conducted a multicenter trial of a MUC1 vaccine in current and former heavy smokers to evaluate immunogenicity and safety. Smoking history of ≥30 pack-years and CT chest showing either no nodules or nodules < 6 mm were inclusion criteria. A vaccine containing MUC1 peptide was administered at weeks 0, 2 and 10. Blood was collected pre-vaccine administration, 2 weeks after each vaccine, and at week 24. Immunogenicity (primary endpoint) and the presence of myeloid-derived suppressor cells (MDSC) and regulatory T cells (secondary endpoints) were assessed. Adverse events and toxicities were monitored.

**Results:** Of 77 individuals screened, 50 were registered and 45 completed the study (27 current and 18 former smokers). The vaccine was well-tolerated. Four current (14.8%) and 2 former smokers (11.1%) developed anti-MUC1 IgG titers ≥2 fold higher at week 12 as compared with baseline, with an overall immune response rate of 13.3% (95% CI 5.1–26.8%). We found high circulating levels of immunosuppressive MDSCs in both current and former smokers.

**Conclusions:** Administering a potentially preventive vaccine is feasible and safe in individuals at high risk for lung cancer. However, this cohort exhibited a high level of immune suppression, previously documented only in patients with advanced lung cancer. Nonetheless, a vaccine-induced immune response was noted in 13% of participants. Further work is required to refine participant selection, understand the factors driving immunosuppression, and counteract these factors to apply immunoprevention strategies in this high-risk population.

**Translational Relevance:** Vaccines targeting antigens aberrantly expressed on lung cancers and their precursors offer the potential for a relatively non-invasive prevention strategy. Worldwide, lung cancer remains the most important cause of cancer-related mortality, and an immunoprevention approach, if successful, has the potential to save many lives by prevention of lung cancer. All epithelial tumors, including lung cancer, express high levels of abnormal mucin (MUC)1 and MUC1-based therapeutic vaccines can increase this immune response to aberrant expressed MUC1 to therapeutic levels.

We conducted a multicenter trial to evaluate the immunogenicity and safety of a MUC1 preventive vaccine in individuals with a significant smoking history and at high risk of developing lung cancer. The MUC1 vaccine was safe and elicited an immune response in 13%, which was lower than anticipated. Unexpectedly, we observed a high level of immune suppression in this cohort of heavy smokers, which has been previously documented in individuals who already have lung cancer. Although immunoprevention strategies are promising for reducing the risk of lung cancer in current and former heavy smokers, the immune environment induced by smoking, and the factors that drive immunosuppression are crucial barriers that must be overcome by well-designed immunoprevention strategies for lung cancer.

## Introduction

In 2024, the American Cancer Society estimated that 234D580 people in the United States would be diagnosed with lung and bronchus cancer, and that 125D070 would die from their disease.(1) Worldwide, an estimated 2.5 million new lung cancer cases were diagnosed in 2022, with an estimated 1.8 million deaths.(2) Tobacco smoking is the most common underlying etiology for lung cancer, with 80–90% of lung cancer patients having a history of smoking.(3, 4) Smoking cessation is an essential component in the prevention of lung cancer. However, even when smoking cessation is successful, risk continues to increase with time albeit at a significantly lower rate when compared with continued smoking.(5, 6) Survival is improving, but it remains poor (25% 5-year survival),(1) and new strategies for prevention of this deadly disease are urgently needed.

In view of the inconsistent results of chemoprevention trials,(7–9) immunoprevention of lung cancer is of increasing interest.(7) We previously demonstrated that the density of tumor-infiltrating lymphocytes correlates with outcomes after resection of stage I non-small cell lung cancer (NSCLC).(10) A higher density of tumor-infiltrating lymphocytes was associated with improved disease-free survival in resected NSCLC ≥ 5 cm. Vaccines targeting antigens aberrantly expressed on lung cancers and their precursors offer the potential for a relatively non-invasive and non-toxic prevention and risk-reduction strategy.

Mucin 1 (MUC1) is a high-molecular-weight transmembrane glycoprotein expressed in normal epithelial cells at low levels, polarized to the apical surface and extensively glycosylated. All epithelial tumors, including lung cancer, express high levels of abnormal MUC1 characterized by hypoglycosylation and loss of luminal polarity.(11) Cancer patients with MUC1-positive tumors produce MUC1-specific antibodies and MUC1-specific T cells, albeit at low levels.(12–14) MUC1-based therapeutic vaccines have been used to increase this immune response to therapeutic levels.(15–18) Immunosuppressive forces in the tumor microenvironment can blunt the potential efficacy of vaccines, however.(19–22) We hypothesized that vaccines administered early, in the absence of cancer but when an individual is at high risk for cancer or during the premalignant phase when immunosuppression is not expected, might generate a more robust immune response.

We have been testing this hypothesis by administering a MUC1 vaccine in individuals likely harboring premalignant disease. The first two trials tested vaccine safety, immunogenicity, and efficacy in individuals at high risk for colon cancer due to a recent history of colonic adenomas.(15, 23) The third study, described here, was a multicenter pilot trial through the NCI-funded Cancer Prevention Network (CPN),to evaluate the immunogenicity and safety of the same MUC1 vaccine in individuals with a significant smoking history and at high risk of developing lung cancer.

## Methods

### Study Design

This was a multicenter study designed for preliminary evaluation of the immunogenicity and safety of a MUC1 vaccine in heavy smokers at high risk for developing lung cancer. All aspects of the study protocol were reviewed and approved by the National Cancer Institute (NCI) Central Institutional Review Board. Participants were enrolled at two sites across two geographic regions of the United States either the University of Pittsburgh/University of Pittsburgh Medical Center (UPMC), Pittsburgh, Pennsylvania or the Mayo Clinic, Rochester, Minnesota. The Cancer Prevention Network at the Mayo Clinic served as the coordinating center. The Data and Safety Monitoring Board of the Mayo Clinic Cancer Institute reviewed the safety data every six months. The study was registered at ClinicalTrials.gov (NCT03300817).

### Eligibility

Prior to screening, all participants provided signed informed consent. The inclusion criteria were based on recommendations for lung cancer screening with CT scans that were current at the time of this study and represented established criteria that characterize individuals as high-risk for the development of lung cancer.(24–27) The study population included males and females, 55–80 years of age, who were current or former smokers with a ≥ 30 pack-year history of smoking. The number of pack years was determined by multiplying the number of packs smoked per day by the number of years smoked. Other inclusion criteria were a chest computerized tomography (CT) scan ≤ 6 months prior to registration indicating no nodules or solid or part-solid nodules < 6 mm in size, consistent with < 1% probability of malignancy as per Lung-RADs Version 1.0; normal organ and marrow function; and Eastern Cooperative Oncology Group (ECOG) performance status ≤ 1. Exclusion criteria included recent history of any malignancy, recent use of investigational agents, prior investigational immune therapy, known HIV or autoimmune diseases, use of systemic steroids, and positive antinuclear antibodies (ANA) titer (with subsequent protocol amendments to define exclusionary ANA titer as > 1:160).

Prior to beginning the intervention, baseline evaluations were performed, including review of medical and surgical history and concomitant medications, a physical exam, and blood tests, lung function with spirometry and urine cotinine testing to verify smoking status. A negative pregnancy test was required for individuals of childbearing potential. All participants completed baseline alcohol and tobacco use assessments.

### MUC1 Vaccine

The vaccine consisted of 100 μg of a 100-amino-acid synthetic MUC1 peptide that included 5 tandem repeats of 20 amino acids. The sequence was derived from the extracellular domain of MUC1, which is known as the variable-number-of-tandem-repeats (VNTR) region.(28) The vaccine peptide H_2_N-(GVTSAPDTRPAPGSTAPPAH)_5_-CONH_2_ was synthesized at the University of Pittsburgh Clinical Peptide Synthesis Facility. Just before injection, it was admixed with Hiltonol® adjuvant (toll-like-receptor-3 agonist, polyinosinic-polycytidylic acid (poly-ICLC,), purchased from Oncovir Inc. (Washington, DC). (29, 30)

### Vaccine protocol and blood sampling

All screened participants deemed eligible to continue to intervention received the MUC1/poly-ICLC vaccine at weeks 0, 2, and 10 with blood testing for safety done prior to each vaccination and at weeks 4, 12, and 24. Blood for immune response assessment was drawn at baseline and at weeks, 2, 4, and 12, with an additional blood draw at week 24 to evaluate durability of the immune response. The vaccine was administered subcutaneously, in the upper arm, and when possible, in the same upper arm at each injection. Participants were monitored for adverse events (AE).

### Primary and secondary endpoints

The primary endpoints were the 12-week immunogenicity with desired immune response defined as an antibody titer that was ≥2 fold higher at 12 weeks as compared with baseline, and the safety of the vaccine assessed according to the National Cancer Institute (NCI) Common Toxicity Criteria (Common Terminology Criteria for Adverse Events (CTCAE)) version (v) 4.0. The final routine post-intervention evaluation of safety was at week 24 and included physical exam and toxicity/AE assessment; repeat lung function spirometry testing; urine cotinine test; post-intervention alcohol and tobacco use assessments; and a quality of life/Was-It-Worth-It questionnaire. Any participants reporting unresolved AEs at week 24 were contacted again at week 28 for further evaluation.

Secondary endpoints included a) exploration of potential differences in the immunogenicity of the vaccine between current and former smokers, and b) evaluation of the pre-vaccination levels of circulating myeloid-derived suppressor cells (MDSC) and potential association with the ability to respond to the vaccine.

### Exploratory endpoints

Exploratory endpoints were: a) to assess duration of the immune response at week 24; b) to explore the relationship between baseline chronic obstructive pulmonary disease (COPD) status and immune response in current vs. former smokers; c) to explore changes in immunogenicity in individuals with COPD with different levels of circulating MDSCs; d) to explore the impact of baseline levels of hsCRP and IL-6 on the ability to successfully vaccinate with MUC1/poly-ICLC and the impact of elicited immunity on IL-6 and hsCRP levels post-vaccination; and e) to establish a biospecimen repository archive containing frozen peripheral blood cells and plasma for future analyses.

### Immune Endpoints

Registered participants who received all 3 vaccinations and completed both the baseline and 12-week post-baseline evaluation were evaluable for the primary study endpoint of immunogenicity of the vaccine. The primary study endpoint was to measure MUC1 IgG levels, which also indirectly measures T-cell-mediated immunity. An anti-MUC1 IgG ratio of ≥2.0 at 12 weeks (week 12/week 0) was defined as a positive immune response, and individuals having an anti-MUC1 IgG ratio of ≥2.0 at 12 weeks were considered vaccine responders. Anti-MUC1 IgG levels were also evaluated at week 24 to measure the stability and durability of the antibody response.

### Adverse Events

AEs and toxicities were closely monitored for up to 24 weeks after participants received the first dose of the MUC1 vaccine. The maximum grade for each type of AE was recorded for each participant and frequency tables were reviewed to determine the overall patterns. In addition, the number and severity of AEs were tabulated and summarized across all grades. Grade ≥3 AEs were similarly described and summarized separately. As per NCI CTCAE v 4.0, toxicities were defined as adverse events that were classified as either possibly, probably, or definitely related to the interventional agent. In addition, we reviewed all AE data and classified the AE as either “related or at least possibly related” or “unrelated or unlikely to be related” to the study intervention.

### Peripheral blood sample collection and processing

Heparinized blood, collected at the University of Pittsburgh or transported there from Mayo Clinic in Rochester, MN, was centrifuged at 2100 rpm for 5 minutes with slow deceleration. The top plasma layer was collected off the top and frozen. The bottom blood layer was diluted 1:1 in Dulbecco’s PBS (DPBS) and layered over 10 ml lymphocyte separation medium (MPbio) in 50ml conical tubes and centrifuged at 800 g for 10 minutes with the lowest acceleration and deceleration speed, within 24 hours from when it was drawn. Peripheral blood mononuclear cells (PBMC) were collected from the interphase, washed twice, re-suspended in 80% human serum and 20% dimethyl sulfoxide (DMSO), and stored in liquid nitrogen at −80°C.

### Anti-MUC1 IgG assay

Enzyme-linked immunosorbent assay (ELISA) was used to measure plasma anti-MUC1 IgG levels, as previously reported. (15) The 100-mer MUC1 vaccine peptide (1 µg/ml) dissolved in 2.5% bovine serum albumin (BSA) in PBS was the capture antigen on Immulon 4HBX ELISA plates (Thermo-Fisher Scientific, MA).

### Myeloid-Derived Suppressor Cells (MDSC) and Treg measurements

Frozen PBMC were thawed in a 37°C water bath, washed, and stained with APC-labeled anti-human CD11b (BD Biosciences Clone:ICRF44), PE-Texas/Red labeled anti-human CD33 (BD Biosciences Clone:WM53), FITC labeled anti-human HLA-DR (BD Biosciences Clone:G46-6), V450 labeled Anti-human CD14 (BD Biosciences Clone:MDP9) and PE-Cy7 labeled anti human CD15 (BD Biosciences Clone:HI98), and anti-S100A (BD Pharmingen) Regulatory T cells were stained with anti-FOXP3and anti-CD25, both from BD Pharmingen). Stained cells were visualized on IMM Fortessa (BD Bioscience) and data analyzed using FlowJo (v10) software (FlowJo LLC).

### Statistical Analysis

The powered primary endpoint was the 12-week immune response defined as MUC1-specific IgG antibody titer ≥2 fold higher at 12 weeks as compared with baseline. A sample size of 40 evaluable participants would provide 96% power to detect an immune response rate of 15% versus 40% using a 2-sided test of proportions with a type I error rate of 0.05. The overall 12-week immune response rate was reported as a percentage with a 95% confidence interval. The other primary endpoint was safety, assessing adverse events (AEs), as described above. AEs were descriptively reported as frequencies and percentages.

For other endpoints, the associations of categorical data were analyzed using the Chi-square test; the association of continuous data with binary data was analyzed with the Wilcoxon-Rank sum test.

## Results

### Study Participants

Seventy-seven participants were pre-registered (screened and consented) over a 3-year period from February 2018 to February 2021, and 50 participants who met the predefined criteria were registered for the trial. The trial was paused for several months due to the COVID-19 pandemic. The CONSORT diagram of participant flow is shown in **Figure 1**. One participant (2%) withdrew prior to starting the intervention and was removed from all analyses, leaving 49 participants, who received at least one vaccine dose. All 49 participants were eligible for assessment of safety and toxicity of vaccine. The median age of the cohort was 67 years (range 56-80 years), with 33 men (67.3%) and 16 women (32.7%), 30 current smokers and 19 former smokers. There were no significant differences in age or sex between the current and former smokers. The participant characteristics are summarized in **Table 1**.

**Figure 1.**
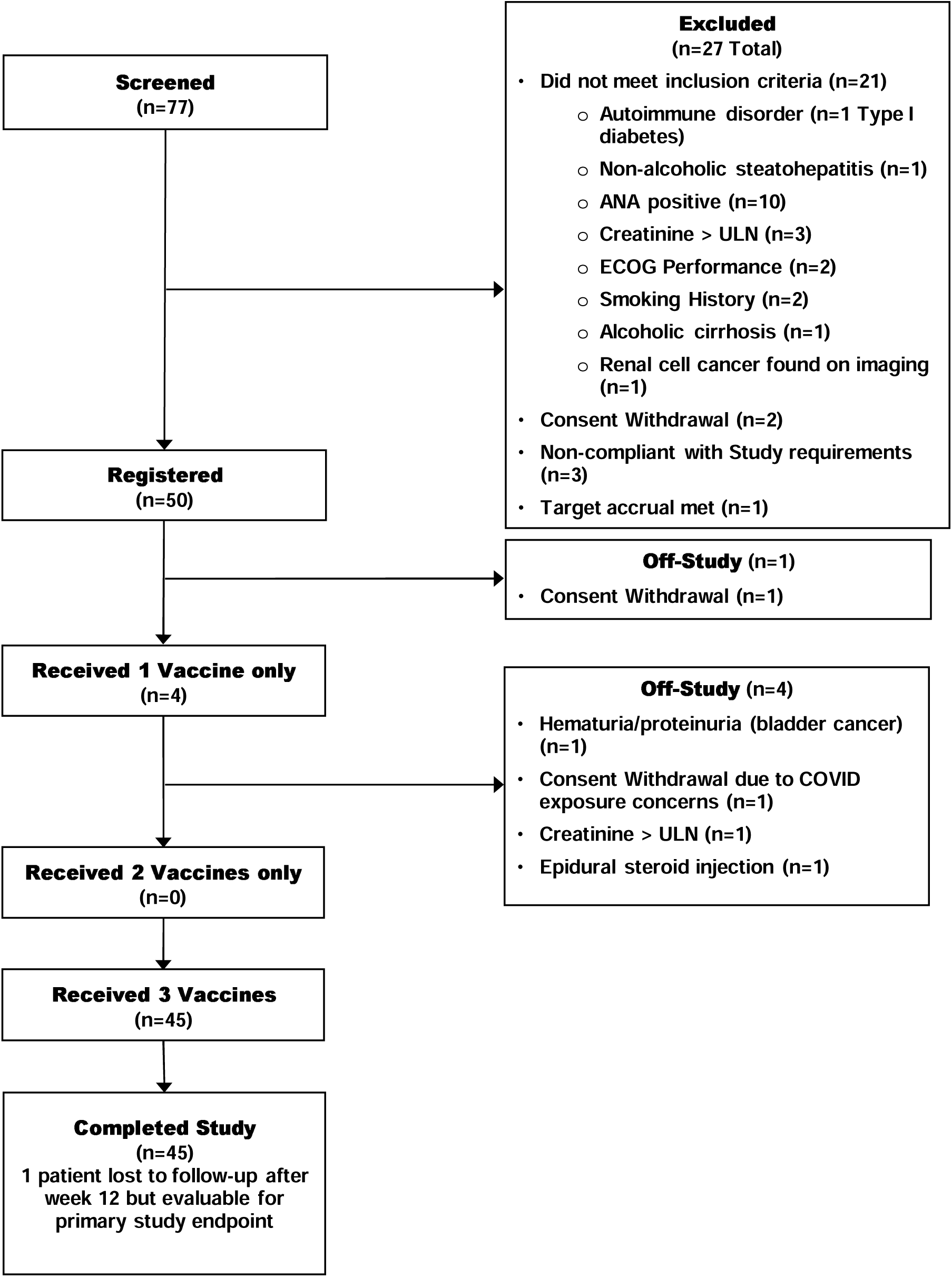
Consort diagram of the pilot study of a MUC1 vaccine in current and former smokers at high risk for lung cancer. ANA, Antinuclear antibody; ECOG, Eastern Cooperative Oncology Group; ULN, upper limit of normal.

**Table 1.**
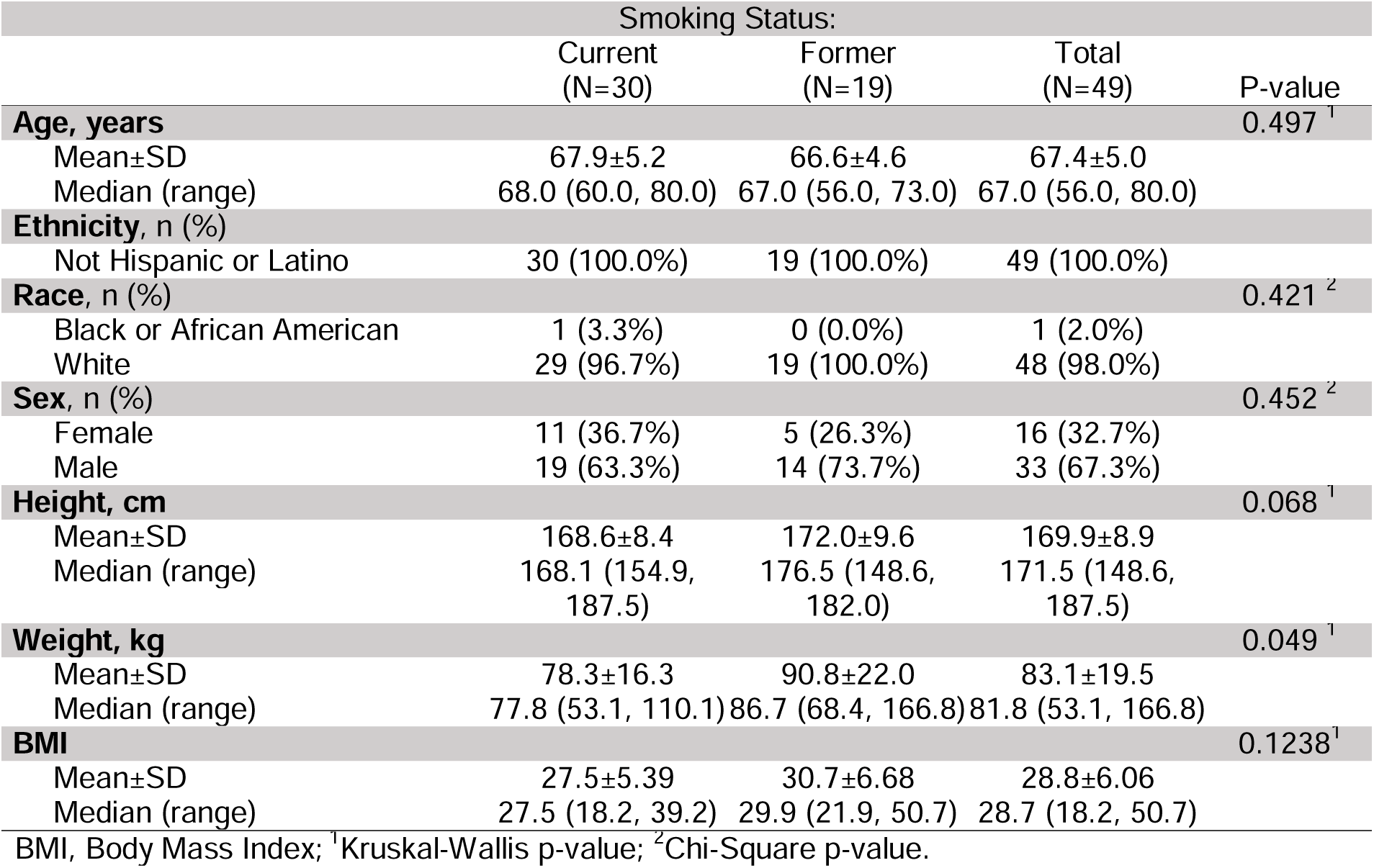
Participant Characteristics.

|  | Smoking Status: |  |  | P-value |
| --- | --- | --- | --- | --- |
|  | Current<br>(N=30) | Former<br>(N=19) | Total<br>(N=49) |  |
| **Age, years** |  |  |  | 0.497 <sup>1</sup> |
| Mean±SD | 67.9±5.2 | 66.6±4.6 | 67.4±5.0 |  |
| Median (range) | 68.0 (60.0, 80.0) | 67.0 (56.0, 73.0) | 67.0 (56.0, 80.0) |  |
| **Ethnicity, n (%)** |  |  |  |  |
| Not Hispanic or Latino | 30 (100.0%) | 19 (100.0%) | 49 (100.0%) |  |
| **Race, n (%)** |  |  |  | 0.421 <sup>2</sup> |
| Black or African American | 1 (3.3%) | 0 (0.0%) | 1 (2.0%) |  |
| White | 29 (96.7%) | 19 (100.0%) | 48 (98.0%) |  |
| **Sex, n (%)** |  |  |  | 0.452 <sup>2</sup> |
| Female | 11 (36.7%) | 5 (26.3%) | 16 (32.7%) |  |
| Male | 19 (63.3%) | 14 (73.7%) | 33 (67.3%) |  |
| **Height, cm** |  |  |  | 0.068 <sup>1</sup> |
| Mean±SD | 168.6±8.4 | 172.0±9.6 | 169.9±8.9 |  |
| Median (range) | 168.1 (154.9, 187.5) | 176.5 (148.6, 182.0) | 171.5 (148.6, 187.5) |  |
| **Weight, kg** |  |  |  | 0.049 <sup>1</sup> |
| Mean±SD | 78.3±16.3 | 90.8±22.0 | 83.1±19.5 |  |
| Median (range) | 77.8 (53.1, 110.1) | 86.7 (68.4, 166.8) | 81.8 (53.1, 166.8) |  |
| **BMI** |  |  |  | 0.1238 <sup>1</sup> |
| Mean±SD | 27.5±5.39 | 30.7±6.68 | 28.8±6.06 |  |
| Median (range) | 27.5 (18.2, 39.2) | 29.9 (21.9, 50.7) | 28.7 (18.2, 50.7) |  |

BMI, Body Mass Index; <sup>1</sup>Kruskal-Wallis p-value; <sup>2</sup>Chi-Square p-value.

All 49 participants received the week 0 vaccine injection; however, 4 participants did not receive any further injections (2 were subsequently deemed ineligible, 1 participant refused follow-up, and 1 experienced AEs unrelated to the vaccine) leaving a total of 45 participants evaluable for response.

### Primary Endpoint: Safety

Six participants experienced grade 3 AEs (12.2%, **Table 2**). Two grade 3 AEs were at least possibly related to the vaccine intervention: one experienced leukocytosis (immune responder) and one, who had a history of alcohol use, experienced a transient increased serum lipase (immune responder). In 4 participants, the AEs were determined not to be related to the vaccine (one participant experienced a fall with multiple rib fractures and a pneumothorax and subsequent hypoxia and respiratory acidosis (non-responder); one experienced hematuria (non-responder), one experienced hyponatremia (non-responder), and one experienced a tooth infection (non-responder). The most prominent grade 2 AE was an injection site reaction (19 participants, 3 responders,16 non-responders), followed by pain (6 participants, all non-responders), and upper respiratory infection (4 participants, 1 responder, 3 non-responders). **Supplemental Table 1** summarizes all the adverse events.

**Table 2.**
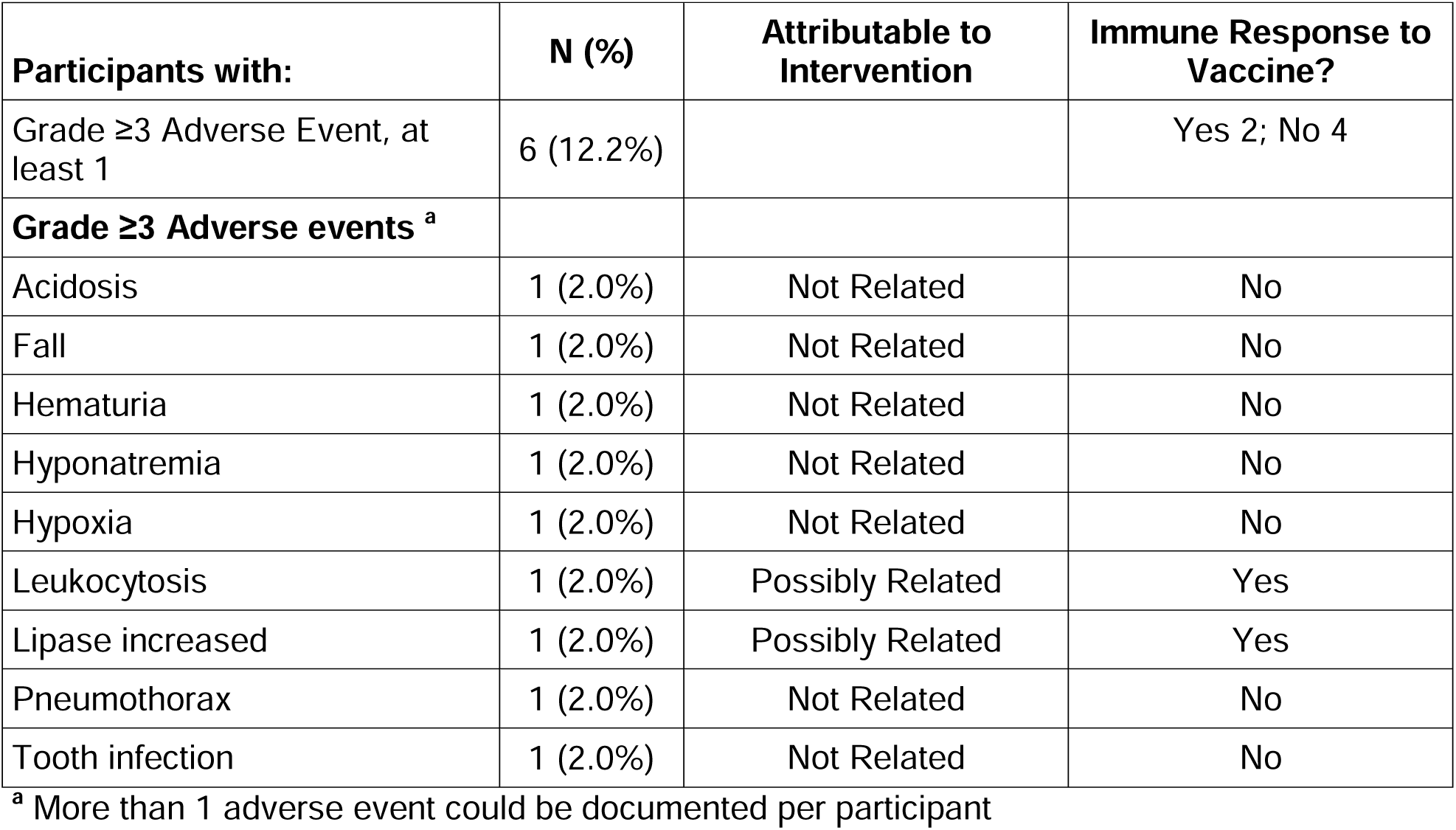
Grade 3 or greater adverse events in 49 evaluable participants.

Of note, the pulmonary status of our study participants (as assessed by physical exams, symptom review throughout the study and the pulmonary function tests done at baseline and week 24) remained stable in 44 of the 45 participants who received 3 doses of the vaccine. No clinically significant worsening of pulmonary function was seen. One participant (a non-responder) had a Grade 1 decrease in forced expiratory volume in 1 second (FEV1) **(Supplemental Table 1)**. Although there were significant differences in pulmonary function between current and former smokers both at baseline and at 24 weeks (**Supplemental Table 2**), there were no significant differences in pulmonary function when comparing immune responders vs. immune non-responders at baseline and at 24 weeks. (**Supplemental Table 3**)

### Primary Endpoint: Vaccine Immunogenicity

There were 6 immune responders to the vaccine (4 of 27 current smokers (14.8%) and 2 of 18 former smokers (11.1%) among the 45 evaluable participants for an overall response rate of 13.3% (95% confidence interval [CI] 5.1%, 26.8%). Pre-vaccination levels of anti-MUC1 antibodies were low in most individuals (**Figure 2**). Three participants had very high anti-MUC1 antibody levels pre-vaccination that did not double after vaccination, and were categorized as non-responders. In 2 immune responders, antibody levels decreased by week 24, but were still higher than baseline, while in the other 4 responders, antibody levels at week 24 remained at the same level as week 12.

**Figure 2.**
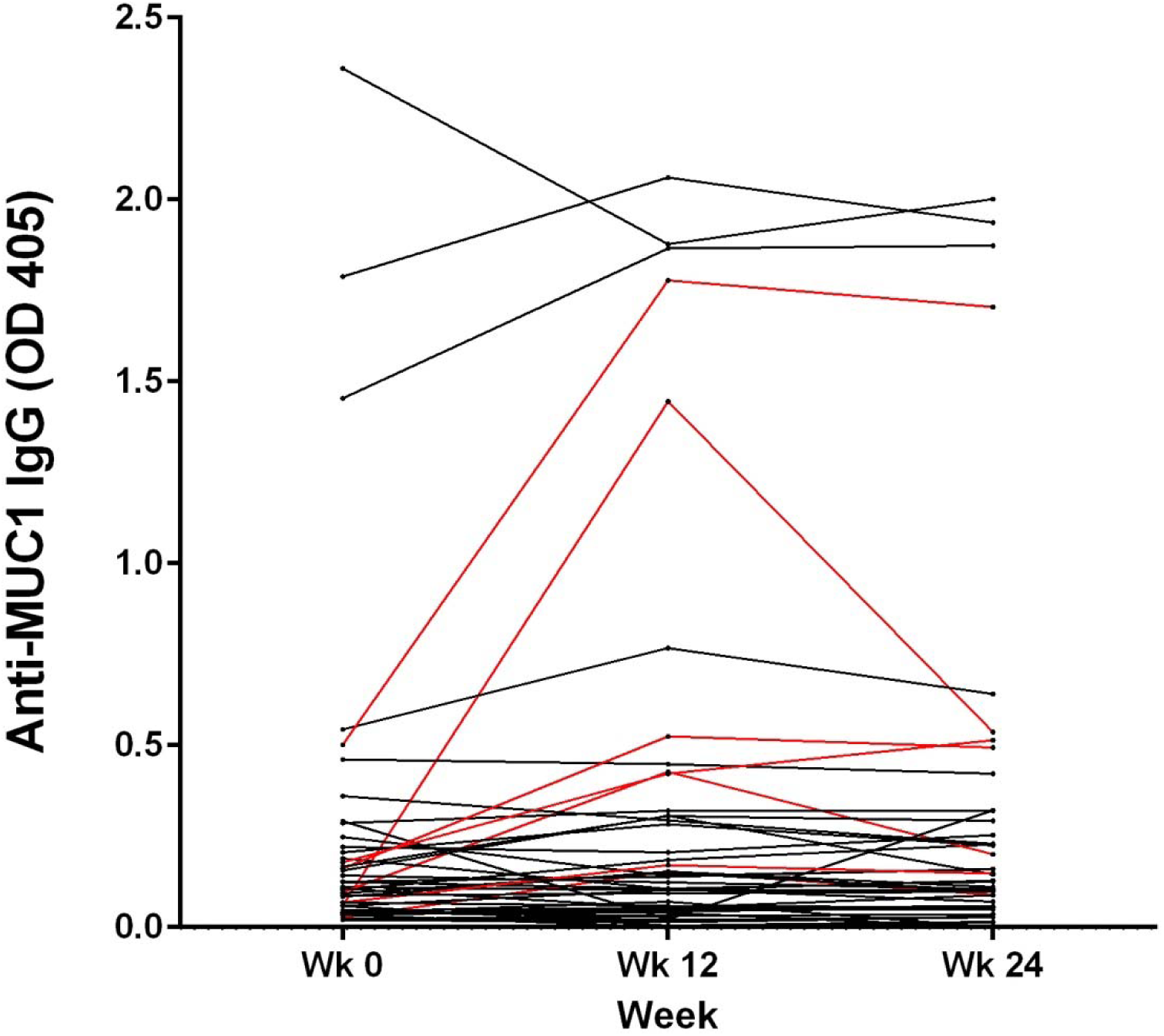
Vaccine-elicited anti-MUC1 antibodies in current and former smokers. Vaccine was given at week 0, 2, and 10 and serum ELISA was performed at weeks 12 and 24. Immune responders (IgG OD ≥2 over baseline) are shown in red and immune non-responders in black.

### Secondary Endpoint: MDSC

Both current and former smokers had high levels of circulating MDSC (median 13.7%; normal range is < 2%)(**Figure 3A**). All MDSC were positive for S100A9 (**Figure 3B**), a marker of functionally immunosuppressive MDSCs.(31) In contrast, and somewhat surprisingly, the levels of circulating T regulatory cells (Treg), characterized by the presence of the transcription factor FoxP3, were within normal ranges (< 2%) in both current and former -smokers (**Figure 3C**).

**Figure 3.**
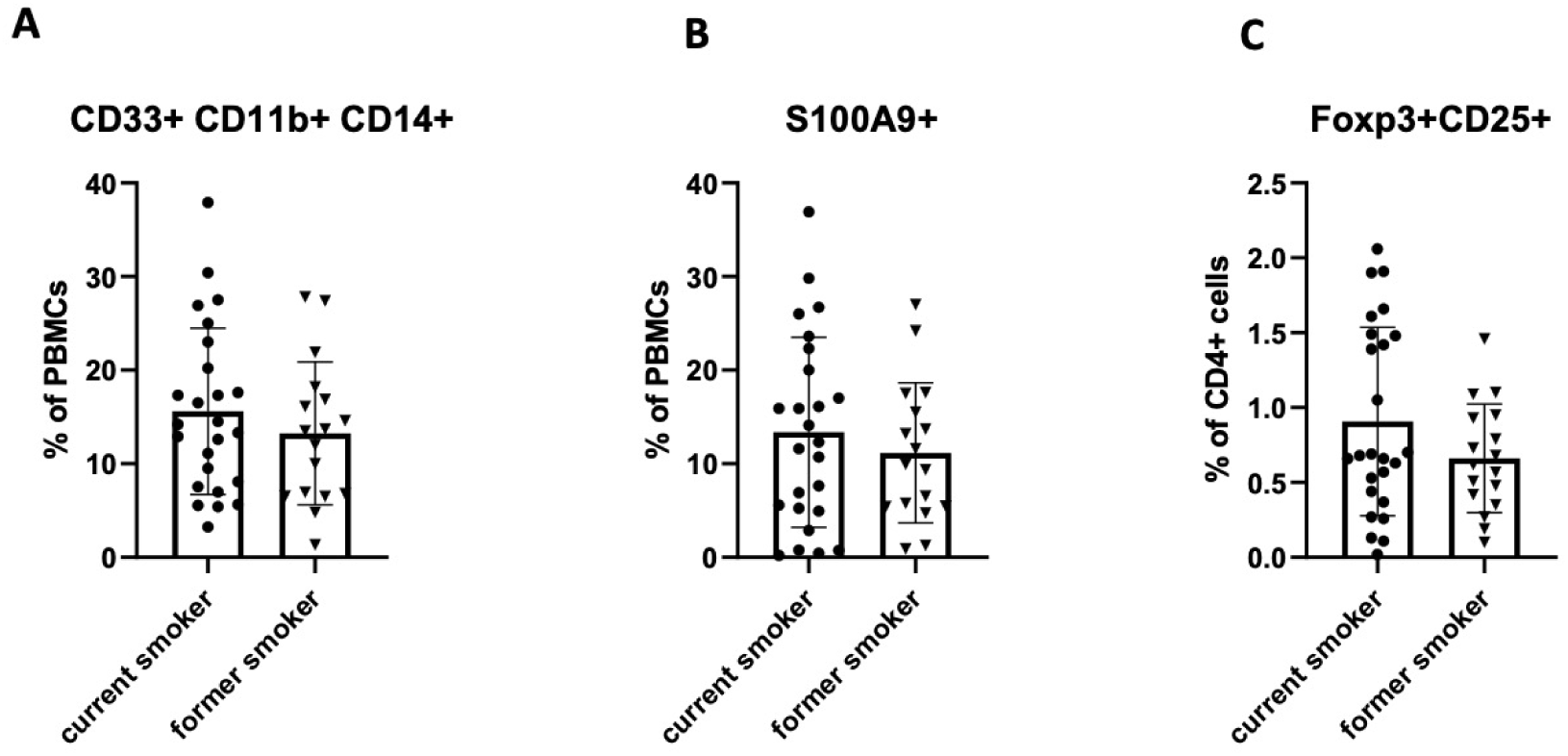
Immunosuppressive cells in peripheral blood mononuclear cells (PBMC) of current smokers and former smokers. **(A)** Myeloid-derived suppressor cells (MDSC) are CD33+CD11b+CD14+; S100A is a marker of functionally immunosuppressive MDSC; **(B)** Regulatory T cells (Treg) are FoxP3+CD25+ **(C)**.

### Exploratory Endpoints

IL-6 and hsCRP levels were not significantly associated with immune response to the vaccine. However, the baseline and week-24 IL-6 levels were significantly associated with the smoking status (p < 0.05) with higher levels observed in current smokers than in former smokers (**Table 3**). Similarly, the baseline IL-6 levels were abnormally elevated in a significantly higher percentage of current smokers (81%) as compared with former smokers (50%, p=0.031). Baseline hsCRP levels were abnormally elevated in 43% of participants, of whom a higher percentage were current smokers (54%) as compared with former smokers (28%, p=0.086) (**Table 4**). Similarly, the median baseline levels of hsCRP were higher in current smokers (0.2 mg/L, range 0.0–1.5 mg/L) as compared with former smokers (0.2 mg/L, range 0.0–1.1 mg/L; p=0.066). At week 24, a significantly elevated median hsCRP level was seen in current smokers (0.2 mg/L, range 0.0–3.1 mg/L) when compared with former smokers (0.1 mg/L, range 0.0–14.8 mg/L; P= 0.016)(**Table 3** and **Table 4**).

**Table 3.** Smoking Status and Inflammation: Circulating IL-6 Levels.

| **IL-6** | **Current Smoker** | **Former Smoker** | **All Participants** | **P-value** |
| --- | --- | --- | --- | --- |
| **IL-6 Baseline, pg/ml** | N=26 | N=18 | N=44 | 0.021 <sup>1</sup> |
| Mean ± SD | 4.3±6.5 | 2.2±1.4 | 3.4±5.1 |  |
| Median (range) | 2.9 (0.4, 35.0) | 1.8 (0.6, 5.4) | 2.3 (0.4, 35.0) |  |
| **IL-6 Baseline status, n (%)** | N=26 | N=18 | N=44 | 0.031 <sup>2</sup> |
| Elevated | 21 (81.0%) | 9 (50.0%) | 30 (68.2%) |  |
| Normal | 5 (19.0%) | 9 (50.0%) | 14 (31.8%) |  |
| **IL-6 Week 24, pg/ml** | N=22 | N=17 | N=39 | 0.001 <sup>1</sup> |
| Mean ± SD | 4.5±4.2 | 3.7±7.1 | 4.1±5.6 |  |
| Median (range) | 3.5 (0.6, 22.0) | 1.8 (0.8, 31.0) | 2.8 (0.6, 31.0) |  |
| **IL-6 Week 24 status, n (%)** | N=22 | N=17 | N=39 | 0.001 <sup>2</sup> |
| Elevated | 21 (95.5%) | 8 (47.1%) | 29 (74.4%) |  |
| Normal | 1 (4.5%) | 9 (52.9%) | 10 (25.6%) |  |

IL, interleukin. <sup>1</sup>Kruskal-Wallis p-value; <sup>2</sup>Chi-Square p-value.

**Table 4.** Smoking Status and Inflammation: Circulating hsCRP Levels.

| hsCRP | Current Smoker | Former smoker | All Participants | P-value |
| --- | --- | --- | --- | --- |
| **hsCRP Baseline, mg/L** | N=26 | N=18 | N=44 | 0.066 <sup>1</sup> |
| **Mean ± SD** | 0.3 ±0.4 | 0.2±0.2 | 0.3±0.3 |  |
| **Median (range)** | 0.2 (0.0, 1.5) | 0.1 (0.0, 1.1) | 0.2 (0.0, 1.5) |  |
| **hsCRP Baseline Status, n (%)** | N=26 | N=18 | N=44 | 0.086 <sup>2</sup> |
| **Elevated** | 14 (53.8%) | 5 (27.8%) | 19 (43.2%) |  |
| **Normal** | 12 (46.2%) | 13 (72.2%) | 25 (56.8%) |  |
| **hsCRP Week 24, mg/L** | N=23 | N=17 | N=40 | 0.016 <sup>1</sup> |
| **Mean ± SD** | 0.5±0.7 | 1.0±3.6 | 0.7±2.4 |  |
| **Median (range)** | 0.2 (0.0, 3.1) | 0.1 (0.0, 14.8) | 0.2 (0.0, 14.8) |  |
| **hsCRP Week 24 Status, n (%)** | N=23 | N=17 | N=40 | 0.066 <sup>2</sup> |
| **Elevated** | 12 (52.2%) | 4 (23.5%) | 16 (40.0%) |  |
| **Normal** | 11 (47.8%) | 13 (76.5%) | 24 (60.0%) |  |

hsCRP, high-sensitivity C-reactive protein. <sup>1</sup>Kruskal-Wallis p-value; <sup>2</sup>Chi-Square p-value.

## Discussion

In this study from two institutions in different regions of the United States, a MUC1 cancer-preventive vaccine was safe with no serious AEs in current and former heavy smokers at high risk for lung cancer. An overall immune response to the vaccine was seen in 13% of participants, with an immune response in 14.8% of the current smokers, and 11.1% of the former smokers. The immune response rate was lower in this cohort of participants at high-risk for lung cancer as compared with previous trials of this vaccine in individuals at risk for colorectal cancer.(15, 23) An elevated level of circulating immunosuppressive S100A+ MDSCs was observed in the majority of study participants and is likely one of the reasons for the lower-than-expected immune response observed. This correlation between MDSC levels in premalignancy and the inability to respond to a vaccine was also seen in MUC1 vaccine trials for colon cancer prevention.(15, 23) The presence of chronic inflammation, which is also highly suppressive to immune responses, was further reflected in elevated levels of serum IL-6 and hsCRP.

Three participants (7%) had high baseline anti-MUC1 antibody levels that did not increase 2-fold post-vaccine, and thus these participants were designated as non-responders to the vaccine even though they were clearly spontaneously responding to abnormal MUC1 expression. One potential reason why these participants did not respond to the vaccine could be that the pre-existing antibodies cleared the vaccine antigen too quickly, thus preventing a boosting effect of the vaccine

The significant immunosuppression observed in our whole study population of heavy smokers was notable, and likely contributed to the lower than anticipated immune response to the study vaccine. The negative impact of smoking on both innate and adaptive immunity has been previously described, and more recently, a few studies have reported an impaired immune response to COVID-19 vaccines in smokers. (32–35) Ferrara and colleagues demonstrated that 60 days after completion of the vaccination cycle with the BioNTech-Pfizer COVID-19 vaccine, serum IgG titers generated in response to the vaccine were significantly lower in current smokers as compared with non-smokers (P=0.002). Nomura and colleagues investigated antibody titers 3 months after the second dose of the same BioNTech-Pfizer COVID-19 vaccine and reported that the antibody titers were, again, significantly lower in current smokers as compared with former smokers (P=0.019).

To better inform the next steps toward immunoprevention in this target population, further work is required to understand the factors driving immunosuppression and the lower-than-expected vaccine-induced immune response in this high-risk population of heavy smokers and to counteract these factors. Additional work is also required for optimal selection of participants who have a higher chance of responding to and potentially benefitting from immunoprevention strategies, including vaccines. Consideration could be given to potential exclusion of subjects with very high baseline levels of circulating MDSCs; however, because the majority of participants in our study had high levels of these immunosuppressive cells, this approach may be of limited value. A more feasible approach could be to combine a cancer vaccine with agents that counter MDSC function, such as pretreatment with high doses of poly-ICLC prior to vaccine administration or in combination with agents that promote T-cell function, such as IL-7 and/or IL-15. (29, 30, 36)

Worldwide, lung cancer remains the most important cause of cancer-related mortality, and an immunoprevention approach, if successful, has the potential to save many lives by prevention of lung cancer.(2) Immunotherapy with PD-1/PD-L1 checkpoint inhibitors improves the survival of patients with lung cancer, and neoadjuvant therapy with PD-1/PD-L1 immunotherapy has become standard of care based on these remarkable findings, despite an objective response rate of only ∼20%.(37, 38) Although we observed a lower than anticipated immune response to the preventive vaccine in this high-risk cohort with a heavy smoking history, if effective, the potential impact of a preventive vaccine worldwide is immense.

### Strengths and Limitations

There were several important strengths of this study. The MUC1 vaccine was shown to be safe in this high-risk population with a heavy smoking history, smoking-associated inflammation, and related comorbidities. The pulmonary functional status of heavy smokers in the study was stable and not adversely affected by the MUC-1 vaccine. The main AEs observed were injection site reactions that are commonly seen after administration of many types of vaccines and resolved without treatment. The study completed its full accrual demonstrating the feasibility of this type of intervention. This was accomplished despite challenges posed by the COVID-19 pandemic and associated restrictions, which necessitated pausing the trial. Moreover, the study was successfully completed in a multi-institutional setting in different geographic regions of the United States.

There were also some significant limitations, and some of these were unexpected. Our study was a small pilot trial designed to evaluate preliminary data on the immune response in a cohort of participants with a heavy smoking history at high risk for lung cancer and to inform potential next steps in the investigation. In addition, the study was powered based on the immune response noted in prior MUC1 vaccine studies in a different population, and we observed a lower-than-anticipated immune response rate as compared with prior studies. We believe this was due to an unexpectedly high rate of significant peripheral immunosuppression in this cohort of heavy smokers. Because of the low numbers of immune responders, we did not have sufficient power to correlate other measurements collected during this study with the immune response. The lower-than-anticipated immune response rate may also have compromised opportunities that the trial could have presented to discover early biomarkers of vaccine responsiveness.

### Conclusions

In summary, in this two-institution pilot study evaluating a MUC1 peptide vaccine in heavy smokers at a high risk for lung cancer, the MUC1 vaccine was safe; however, immune response to the vaccine was lower than anticipated, when compared with that seen in individuals at risk for colon cancer. The observation of a high level of immune suppression in this cohort of heavy smokers, which has been previously documented in individuals who already have lung cancer, is of great interest, however. This demonstrates the need for further research to understand the immune regulation of lung cancer development, the immune environment induced by smoking within lungs and the periphery, and the factors that drive immunosuppression and progression of lung carcinogenesis in this population. Increasing our understanding in these areas will lead to well-designed immunoprevention strategies for lung cancer.

## Data Availability

All data produced in the present study are available upon reasonable request to the authors

## Abbreviations

AE: Adverse event
BSA: Bovine serum albumin
CT: Computed tomography
CTCAE: Common Terminology Criteria for Adverse Events
ECOG: Eastern Cooperative Oncology Group
ELISA: Enzyme-linked immunosorbent assay
FEV1: Forced expiratory volume in 1 second
hsCRP: High sensitivity C-reactive protein
IL: Interleukin
MDSC: Myeloid-derived suppressor cells
MUC1: Mucin 1
OD: Optical density
PBMC: Peripheral blood mononuclear cells
Poly-ICLC: Polyinosinic-polycytidylic acid
Treg: Regulatory T cells

## Acknowledgements

This work was supported by NIH/NCI contract HHSN261201200042I. Dr. Shannon Wyszomierski PhD, assisted in preparing the manuscript for submission with funding from the Sampson Family Endowed Chair in Thoracic Surgical Oncology at the University of Pittsburgh (held by Arjun Pennathur, MD).

## Contributions

**O.J. Finn and A. Pennathur:** Conceptualization, supervision, resources, funding acquisition, investigation, methodology, writing–original draft, project administration, writing–review and editing.

**J. Ward, J. Forster, T. Krpata and S. Fatis:** Investigation, project administration.

**J. McKolanis, J. Xue., P. Beatty:** Investigation.

**S.F. Kaufman, A. Holland, and L. Ambulay:** Project administration.

**C. Akerley, A. Felt and K. Fursa:** Investigation, writing–review and editing.

**M. Wojtowicz, E. Szabo and L. Bengtson:** Conceptualization, supervision, project administration, writing–review and editing.

**N.R. Foster and C. Strand:** Formal analysis, supervision, writing–review and editing.

**A. Salazar:** Resources, writing-review, editing.

**P.J. Limburg and D. Midthun:** Conceptualization, supervision, investigation, methodology, writing–review editing.

## Declarations of Interest

Dr. Limburg serves as Chief Medical Officer for Screening at Exact Sciences through a contracted services agreement with Mayo Clinic. Dr. Limburg and Mayo Clinic have contractual rights to receive royalties through this agreement. Dr. Finn is on the external advisory boards of PDS Biotech, GeoVax and Invectys.

## Data Sharing

Deidentified participant data and supporting trial study documents can be made available upon reasonable request after publication. Data sharing requests should be directed to the corresponding authors.

## Supplementary Material

A Pilot Study of MUC1 Vaccine in Current and Former Smokers at High Risk for Lung Cancer Olivera J. Finn, PhD et al.

**Supplemental Table 1.** Adverse events (all grades), regardless of attribution *N= 49 evaluable participants*.

| Adverse Event | AE grade |  |  |
| --- | --- | --- | --- |
|  | 1 | 2 | 3 |
|  | N (%) | N (%) | N (%) |
| Acidosis |  |  | 1 (2.0%) |
| Acute kidney injury | 1 (2.0%) |  |  |
| Alkaline phosphatase increased | 1 (2.0%) |  |  |
| Allergic reaction | 1 (2.0%) |  |  |
| Anorexia | 1 (2.0%) |  |  |
| Atelectasis |  | 1 (2.0%) |  |
| Bruising |  | 1 (2.0%) |  |
| Chronic kidney disease |  | 1 (2.0%) |  |
| Cough | 1 (2.0%) |  |  |
| Creatinine increased | 1 (2.0%) |  |  |
| Dizziness | 1 (2.0%) | 1 (2.0%) |  |
| Dyspepsia |  | 1 (2.0%) |  |
| Edema face |  | 1 (2.0%) |  |
| Erythema multiforme | 1 (2.0%) |  |  |
| Eye disorders - Other, specify | 1 (2.0%) | 1 (2.0%) |  |
| Fall |  | 1 (2.0%) | 1 (2.0%) |
| Flu like symptoms | 1 (2.0%) |  |  |
| Forced expiratory volume decreased | 1 (2.0%) |  |  |
| General disorders and administration site conditions - Other, specify | 3 (6.1%) | 2 (4.1%) |  |
| Headache | 3 (6.1%) | 1 (2.0%) |  |
| Hematuria |  |  | 1 (2.0%) |
| Hyponatremia | 1 (2.0%) |  | 1 (2.0%) |
| Hypoxia |  |  | 1 (2.0%) |
| Immune system disorders - Other, specify | 1 (2.0%) |  |  |
| Infections and infestations - Other, specify | 2 (4.1%) |  |  |
| Injection site reaction | 18 (36.7%) | 19 (38.8%) |  |
| Injury, poisoning and procedural complications - Other, specify | 1 (2.0%) |  |  |
| Investigations - Other, specify | 3 (6.1%) | 1 (2.0%) |  |
| Leukocytosis |  |  | 1 (2.0%) |
| Lipase increased | 1 (2.0%) | 3 (6.1%) | 1 (2.0%) |
| Muscle weakness upper limb | 1 (2.0%) |  |  |
| Nasal congestion | 1 (2.0%) |  |  |
| Neck pain | 1 (2.0%) |  |  |
| Non-cardiac chest pain |  | 1 (2.0%) |  |
| Pain | 1 (2.0%) | 6 (12.2%) |  |
| Pain in extremity | 1 (2.0%) |  |  |
|  | 1 | 2 | 3 |
|  | N (%) | N (%) | N (%) |
| Pneumothorax |  |  | 1 (2.0%) |
| Proteinuria |  | 1 (2.0%) |  |
| Pruritus | 1 (2.0%) |  |  |
| Rash maculo-papular | 1 (2.0%) |  |  |
| Renal and urinary disorders - Other, specify | 1 (2.0%) |  |  |
| Respiratory, thoracic and mediastinal disorders - Other, specify | 1 (2.0%) | 1 (2.0%) |  |
| Serum amylase increased | 3 (6.1%) | 2 (4.1%) |  |
| Sinus disorder | 1 (2.0%) |  |  |
| Skin and subcutaneous tissue disorders - Other, specify | 1 (2.0%) |  |  |
| Skin infection |  | 1 (2.0%) |  |
| Surgical and medical procedures - Other, specify |  | 3 (6.1%) |  |
| Tooth infection |  |  | 1 (2.0%) |
| Transient ischemic attacks | 1 (2.0%) |  |  |
| Upper respiratory infection |  | 4 (8.2%) |  |
| Urinary tract infection |  | 1 (2.0%) |  |
| Wound dehiscence | 1 (2.0%) |  |  |

**Supplemental Table 2.**
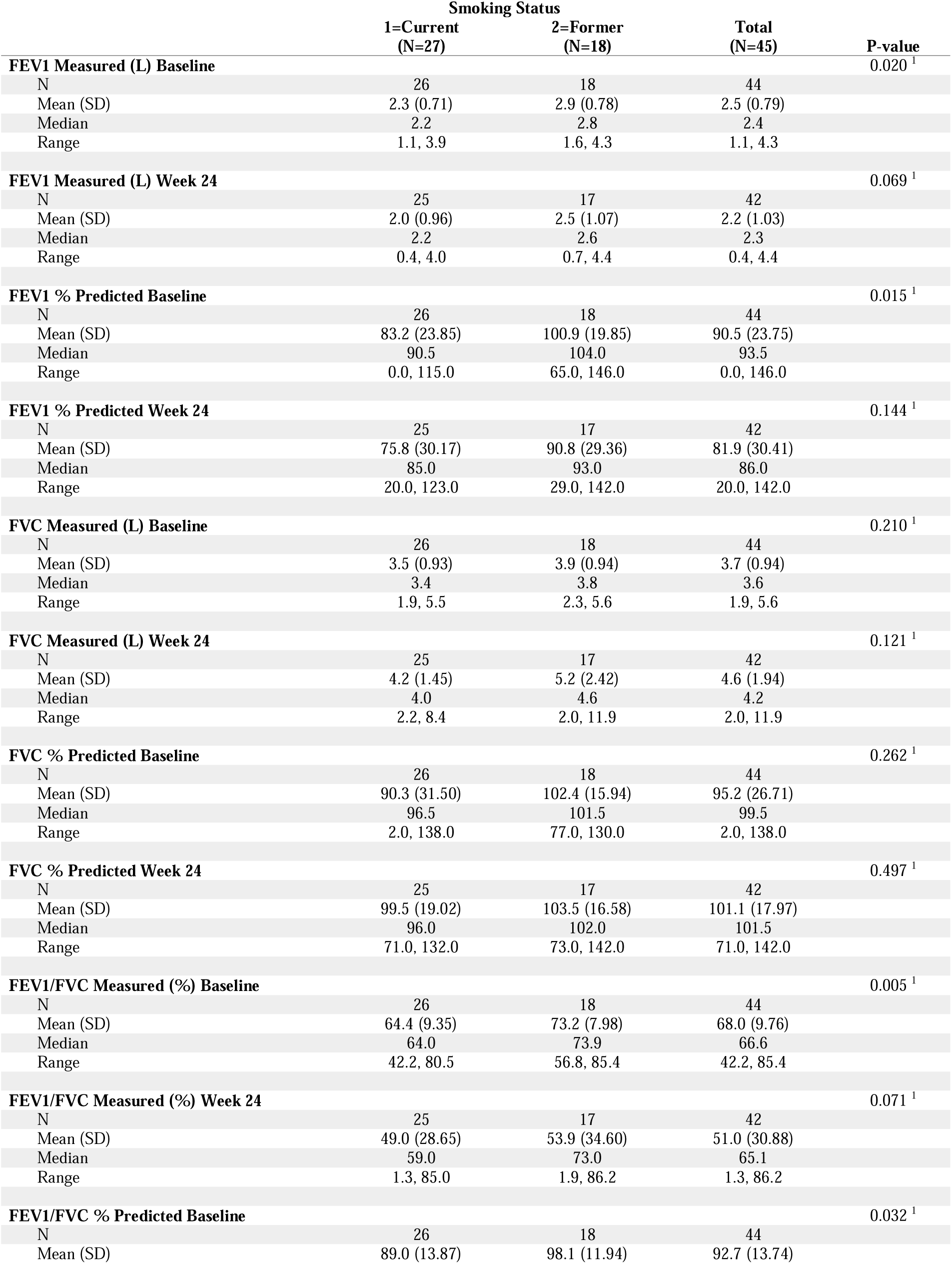
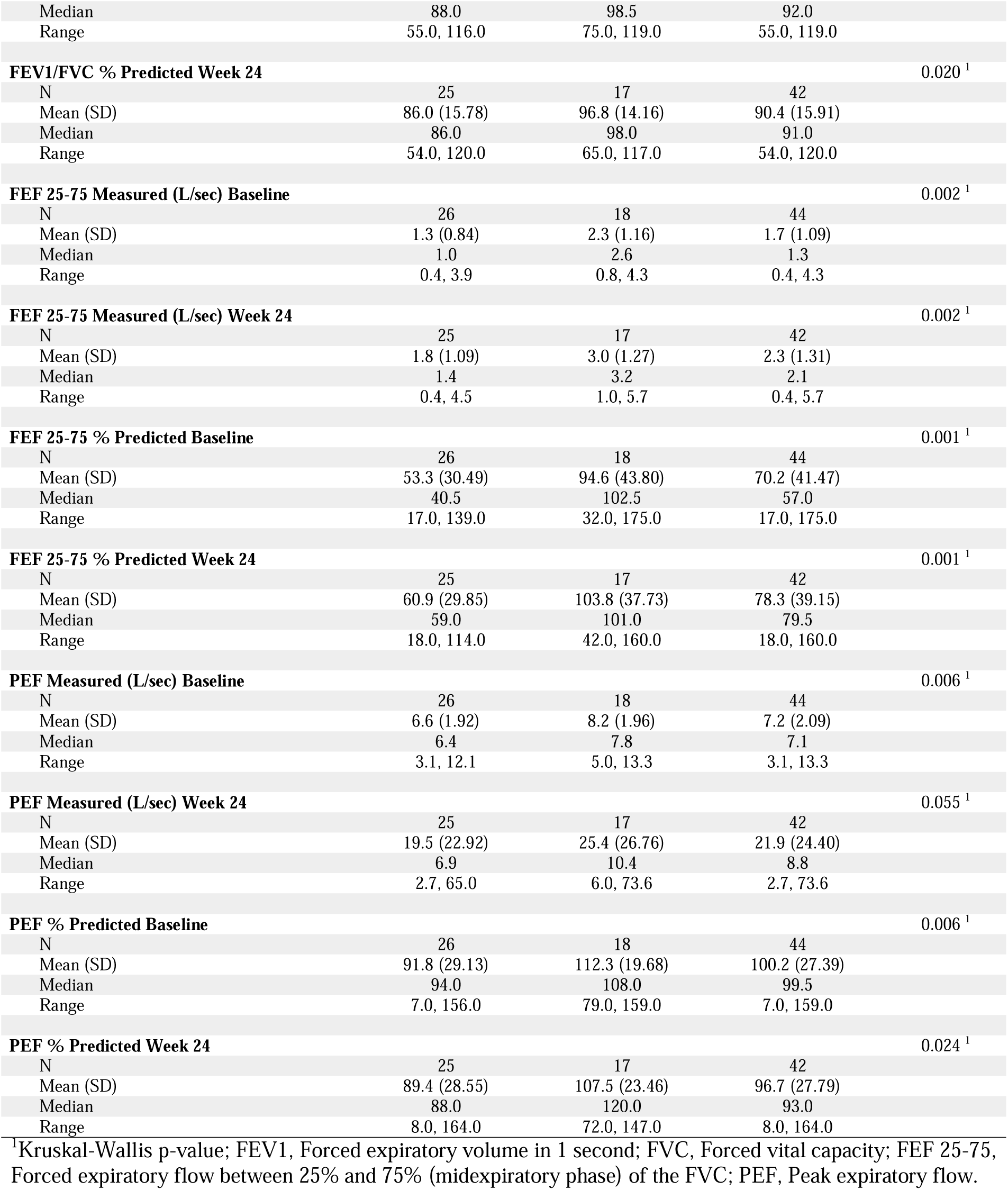
Pulmonary Function Test Results at Baseline and at Week 24 Stratified by Current Smoker vs. Former Smoker.

**Supplemental Table 3.** Pulmonary Function Test Results at Baseline and at Week 24 Stratified by Immune Response.

|  | Immune Non-Responder<br>(N=39) | Immune Responder<br>(N=6) | Total<br>(N=45) | P-value |
| --- | --- | --- | --- | --- |
| **FEV1 Measured (L) Baseline** |  |  |  | 0.905<sup>1</sup> |
| N | 38 | 6 | 44 |  |
| Mean (SD) | 2.5 (0.80) | 2.5 (0.72) | 2.5 (0.79) |  |
| Median | 2.4 | 2.4 | 2.4 |  |
| Range | 1.1, 4.3 | 1.5, 3.7 | 1.1, 4.3 |  |
| **FEV1 Measured (L) Week 24** |  |  |  | 0.472<sup>1</sup> |
| N | 36 | 6 | 42 |  |
| Mean (SD) | 2.2 (1.06) | 2.4 (0.86) | 2.2 (1.03) |  |
| Median | 2.3 | 2.5 | 2.3 |  |
| Range | 0.4, 4.4 | 0.9, 3.6 | 0.4, 4.4 |  |
| **FEV1 % Predicted Baseline** |  |  |  | 0.245<sup>1</sup> |
| N | 38 | 6 | 44 |  |
| Mean (SD) | 91.8 (24.05) | 82.2 (21.83) | 90.5 (23.75) |  |
| Median | 94.5 | 77.0 | 93.5 |  |
| Range | 0.0, 146.0 | 55.0, 114.0 | 0.0, 146.0 |  |
| **FEV1 % Predicted Week 24** |  |  |  | 0.746<sup>1</sup> |
| N | 36 | 6 | 42 |  |
| Mean (SD) | 82.2 (31.27) | 80.0 (27.11) | 81.9 (30.41) |  |
| Median | 87.5 | 82.0 | 86.0 |  |
| Range | 20.0, 142.0 | 33.0, 112.0 | 20.0, 142.0 |  |
| **FVC Measured (L) Baseline** |  |  |  | 0.864<sup>1</sup> |
| N | 38 | 6 | 44 |  |
| Mean (SD) | 3.7 (0.94) | 3.8 (1.03) | 3.7 (0.94) |  |
| Median | 3.6 | 3.5 | 3.6 |  |
| Range | 1.9, 5.6 | 2.6, 5.5 | 1.9, 5.6 |  |
| **FVC Measured (L) Week 24** |  |  |  | 0.914<sup>1</sup> |
| N | 36 | 6 | 42 |  |
| Mean (SD) | 4.6 (2.04) | 4.5 (1.30) | 4.6 (1.94) |  |
| Median | 4.2 | 4.4 | 4.2 |  |
| Range | 2.0, 11.9 | 3.0, 6.2 | 2.0, 11.9 |  |
| **FVC % Predicted Baseline** |  |  |  | 0.732<sup>1</sup> |
| N | 38 | 6 | 44 |  |
| Mean (SD) | 95.5 (27.92) | 93.3 (19.04) | 95.2 (26.71) |  |
| Median | 99.5 | 95.0 | 99.5 |  |
| Range | 2.0, 138.0 | 65.0, 115.0 | 2.0, 138.0 |  |
| **FVC % Predicted Week 24** |  |  |  | 0.787<sup>1</sup> |
| N | 36 | 6 | 42 |  |
| Mean (SD) | 101.4 (17.98) | 99.5 (19.59) | 101.1 (17.97) |  |
| Median | 101.5 | 94.5 | 101.5 |  |
| Range | 71.0, 142.0 | 81.0, 129.0 | 71.0, 142.0 |  |
| **FEV1/FVC Measured (%) Baseline** |  |  |  | 0.632<sup>1</sup> |
| N | 38 | 6 | 44 |  |
| Mean (SD) | 68.4 (9.65) | 65.8 (11.10) | 68.0 (9.76) |  |
| Median | 68.9 | 66.5 | 66.6 |  |
| Range | 42.2, 85.4 | 51.3, 84.7 | 42.2, 85.4 |  |
| **FEV1/FVC Measured (%) Week 24** |  |  |  | 1.000<sup>1</sup> |
| N | 36 | 6 | 42 |  |
| Mean (SD) | 50.2 (31.49) | 55.9 (29.09) | 51.0 (30.88) |  |
| Median | 63.4 | 65.9 | 65.1 |  |
| Range | 1.3, 85.0 | 1.9, 86.2 | 1.3, 86.2 |  |
| **FEV1/FVC % Predicted Baseline** |  |  |  | 0.472<sup>1</sup> |
| N | 38 | 6 | 44 |  |
| Mean (SD) | 93.4 (13.81) | 88.2 (13.57) | 92.7 (13.74) |  |
| Median | 93.5 | 87.0 | 92.0 |  |
| Range | 55.0, 119.0 | 66.0, 106.0 | 55.0, 119.0 |  |
| **FEV1/FVC % Predicted Week 24** |  |  |  | 0.517 <sup>1</sup> |
| N | 36 | 6 | 42 |  |
| Mean (SD) | 91.3 (15.76) | 85.2 (17.28) | 90.4 (15.91) |  |
| Median | 93.5 | 87.0 | 91.0 |  |
| Range | 54.0, 120.0 | 62.0, 108.0 | 54.0, 120.0 |  |
| **FEF 25-75 Measured (L/sec) Baseline** |  |  |  | 0.837 <sup>1</sup> |
| N | 38 | 6 | 44 |  |
| Mean (SD) | 1.7 (1.11) | 1.5 (0.99) | 1.7 (1.09) |  |
| Median | 1.3 | 1.3 | 1.3 |  |
| Range | 0.4, 4.3 | 0.5, 3.3 | 0.4, 4.3 |  |
| **FEF 25-75 Measured (L/sec) Week 24** |  |  |  | 0.565 <sup>1</sup> |
| N | 36 | 6 | 42 |  |
| Mean (SD) | 2.3 (1.35) | 1.9 (1.05) | 2.3 (1.31) |  |
| Median | 2.3 | 1.5 | 2.1 |  |
| Range | 0.4, 5.7 | 0.8, 3.4 | 0.4, 5.7 |  |
| **FEF 25-75 % Predicted Baseline** |  |  |  | 0.452 <sup>1</sup> |
| N | 38 | 6 | 44 |  |
| Mean (SD) | 71.8 (41.13) | 60.2 (46.18) | 70.2 (41.47) |  |
| Median | 59.0 | 48.0 | 57.0 |  |
| Range | 20.0, 175.0 | 17.0, 148.0 | 17.0, 175.0 |  |
| **FEF 25-75 % Predicted Week 24** |  |  |  | 0.429 <sup>1</sup> |
| N | 36 | 6 | 42 |  |
| Mean (SD) | 79.5 (38.79) | 70.7 (44.20) | 78.3 (39.15) |  |
| Median | 84.0 | 56.0 | 79.5 |  |
| Range | 18.0, 160.0 | 32.0, 156.0 | 18.0, 160.0 |  |
| **PEF Measured (L/sec) Baseline** |  |  |  | 0.784 <sup>1</sup> |
| N | 38 | 6 | 44 |  |
| Mean (SD) | 7.3 (2.16) | 7.1 (1.69) | 7.2 (2.09) |  |
| Median | 7.2 | 6.6 | 7.1 |  |
| Range | 3.1, 13.3 | 5.0, 9.5 | 3.1, 13.3 |  |
| **PEF Measured (L/sec) Week 24** |  |  |  | 0.829 <sup>1</sup> |
| N | 36 | 6 | 42 |  |
| Mean (SD) | 22.8 (24.88) | 16.8 (22.69) | 21.9 (24.40) |  |
| Median | 9.2 | 7.2 | 8.8 |  |
| Range | 2.7, 73.6 | 6.1, 63.0 | 2.7, 73.6 |  |
| **PEF % Predicted Baseline** |  |  |  | 0.281 <sup>1</sup> |
| N | 38 | 6 | 44 |  |
| Mean (SD) | 101.3 (28.37) | 93.0 (20.56) | 100.2 (27.39) |  |
| Median | 100.5 | 90.5 | 99.5 |  |
| Range | 7.0, 159.0 | 67.0, 120.0 | 7.0, 159.0 |  |
| **PEF % Predicted Week 24** |  |  |  | 0.774 <sup>1</sup> |
| N | 36 | 6 | 42 |  |
| Mean (SD) | 96.4 (28.97) | 98.5 (21.32) | 96.7 (27.79) |  |
| Median | 94.0 | 91.5 | 93.0 |  |
| Range | 8.0, 164.0 | 77.0, 126.0 | 8.0, 164.0 |  |

<sup>1</sup>Kruskal-Wallis p-value; FEV1, Forced expiratory volume in 1 second; FVC, Forced vital capacity; FEF 25-75, Forced expiratory flow between 25% and 75% (midexpiratory phase) of the FVC; PEF, Peak expiratory flow.

## Notes

**Conflict of Interest Disclosures:** Dr. Limburg serves as Chief Medical Officer for Screening at Exact Sciences through a contracted services agreement with Mayo Clinic. Dr. Limburg and Mayo Clinic have contractual rights to receive royalties through this agreement. Dr. Finn is on the external advisory boards of PDS Biotech, GeoVax and Invectys.

**Funding**: Supported by the National Cancer Institute, contract HHSN261201200042I.<colcnt=2>

### Clinical Trial

NCT03300817

### Funding Statement

This study was funded by the National Cancer Institute, contract HHSN261201200042I.

### Author Declarations

All aspects of the study protocol were reviewed and approved by the National Cancer Institute Central Institutional Review Board.

